# Chest MRI of patients with COVID-19: a retrospective case study

**DOI:** 10.1101/2020.05.13.20088732

**Authors:** Yu.A. Vasilev, K.A. Sergunova, A.V. Bazhin, A.G. Masri, Yu.N. Vasileva, D.S. Semenov, N.D. Kudryavtsev, O.Yu. Panina, A.N. Khoruzhaya, V.V. Zinchenko, E.S. Akhmad, A.V. Petraikin, A.V. Vladzymyrskyy, A.V. Midaev, S.P. Morozov

## Abstract

During the pandemic of COVID-19, computed tomography showed its effectiveness in diagnosis of coronavirus infection. However, ionizing radiation during CT studies causes concern for patients who require dynamic observation, as well as for examination of children and young people. For this retrospective study, we included 15 suspected for COVID-19 patients who were hospitalized in April 2020, Russia. There were 4 adults with positive polymerase chain reaction (PCR) test for COVID-19. All patients underwent MRI examinations using MR-LUND PROTOCOL: Single-shot Fast Spin Echo (SSFSE), LAVA 3D and IDEAL 3D, EPI diffusion-weighted imaging (DWI) and Fast Spin Echo (FSE) T2WI without using respiratory or any other trigger. 3 patients also had CT scan performed. In 5 (33,3%) patients, detected lesioins were visualized on T2WI and DWI simultaneously. At the same time, 4 (26.7%) patients revealed lung tissue changes only on T2WI (P(McNemar)= 0,125, OR= 0,00 (95%), kappa=0,500). Pulmonary changes on MRI were also analyzed depending on their localization. In those patients who had CT scan, the changes were comparable to MRI. MRI of the lungs can detect features of viral pneumonia and assess severity of lung damage. The method can be used to diagnose COVID-19. These data may be applicable for interpreting other studies, such as thoracic spine MRI, detecting signs of viral pneumonia of asymptomatic patients. The current study was limited by a small sample size and absence of chest CT scans of all patients.

## 1. Introduction

The novel coronavirus disease (COVID-19) in some cases is complicated by severe damage of the respiratory system. Recommendations on diagnostics and treatment of suspected for viral pneumonia patients require chest computed tomography (CT).

However, CT scans may not be available due to increased workload on CT rooms during a pandemic. Unfortunately, X-ray and sonography used in such cases do not allow to estimate a volume of lung lesions; therefore, alternative diagnostic methods are required.

The purpose of this study was to evaluate the possibility of using chest MRI as the alternative, non-ionizing method for patients with viral pneumonia. Applying this method, among others, for monitoring COVID-19 patients during and after therapy may increase overall effectiveness of the radiology department.

## 2. Materials and Methods

### 2.1. Study design and participants

This retrospective observational study was approved by the Ethics Committee for the Protection of Human Subjects of Morozov Children University Hospital of Moscow Healthcare Department (Approval №. 156). This study includes a series of 15 cases performed at the Hospital of Chechen Republic, Russia in April, 2020. Age of patients ranged from 26 to 69 years; the average age was 41 ± 13 years. 9 (60%) patients had cold-like symptoms. Also, 2 (13%) patients had cough, shortness of breath, and chest pain. Two patients (13%) did not have any complaints. 4 (26.7%) patients had SARS-CoV-2-positive results of polymerase chain reaction (PCR) test. All these 4 patients were medical employees of the intensive care unit at the infectious diseases department.

### 2.2. MRI scan

All patients underwent chest MRI scan in accordance with the concept “less scan time, more information”. The study was conducted on 3T scanner (Signa Pioneer, GE) in the supine position using the abdominal and spinal RF coils. The center of the abdominal coil was positioned at the mid-sternum. Scans performed without using a respiratory trigger and without holding a breath, when it was possible.

First of all, T2-weighted images (T2WI) were obtained in three planes using a Single-shot Fast Spin Echo (SSFSE) with following parameters: TR 2339.3 ms, TE 89 ms, flip angle 90°, FOV 450×450 mm, matrix (frequency × phase) 384×256, slice thickness 6 mm, spacing between slices 6 mm, number of averages 0.6, k-space filling method – Cartesian. Then these images were applied for planning axial Fast Spin Echo (FSE) T2WI.

T1WI were performed by LAVA 3D and IDEAL 3D. For LAVA 3D, scan parameters were TR 4 ms, TE 2.2 ms and 1,1 ms, flip angle 10°, FOV 400×400 mm, matrix 288×288, slice thickness 3 mm, spacing between slices 1,5 mm, number of averages 0.7 with WATER and FAT fractions, in-phase/out-phase. For IDEAL 3D, scan parameters were TR 5,8 ms, TE 2.5 ms, flip angle 3°, FOV 440×440 mm, matrix 256×256, slice thickness 10 mm, spacing between slices 10 mm, number of averages 0.7 with WATER and FAT fractions, in-phase/out-phase.

Diffusion-weighted imaging (DWI) was performed by EPI pulse sequence TR 10000 ms, TE 62.3 ms, flip angle 89°, FOV 400×400 mm, matrix 128×140, slice thickness 5 mm, spacing between slices 5 mm, number of averages 1, b-values: 50, 800 sec/mm^2^, no respiratory synchronization was used.

In addition, Fast Spin Echo (FSE) T2WI were obtained in axial plane with following parameters: TR 2125 ms, TE 82.7 ms, flip angle 111°, FOV 400×400 mm, matrix 448×256, slice thickness 5 mm, spacing between slices 5 mm, number of averages 2, R–L phase encoding. The scan was divided into parts (covers, batches) to collect data from separate blocks of slices and reduce movement artifacts.

The use of respiratory trigger was complicated by dyspnea in patients with viral pneumonia. Moreover, using respiratory gating for synchronizing MR data collection with respiration requires additional time. In this case, the number of averages for SSFSE, LAVA-Flex and EPI series were chosen not more than 1 in order to reduce blur artifacts associated with non-availability of respiratory gating.

In cases, where differential diagnosis of ground-glass opacity (GGO) is required, breath-hold scans should be conducted at the end of exhalation as well as at the full inhalation. Therefore, during inhalation the parenchyma density decreased due to preservation of partial pneumatization, and the intensity of the MR signal in areas of GGO decreased.

### 2.3. CT scan

Three patients underwent a CT scan after MRI (in less than two days). We used a 128-slice CT scanner (Revolution EVO, GE) and a standard clinical protocol: 120 kV, with adaptive tube current modulation, exposure time 400 ms, slice thickness 1.25 mm, spaces between slices 0.5 mm, LUNG reconstruction filter. The average effective dose was 2 mSv.

### 2.4. Radiologic assessment

Patient studies were analyzed by 5 radiologists with more than 10 years of experience in interpreting MR-images. All safety measures have been followed [17]. In the process of writing a study radiology report, the attention was paid to the presence of polysegmented sections of the isointense signal corresponding to GGO.

Images were also evaluated for the presence of areas of homogeneous hyperintense signal corresponding to pulmonary consolidation on CT scans, as well as for ‘crazy-paving’ sign, which reflects a combination of GGO and pronounced thickening of interlobular septa interstitium.

### 2.5. Statistical analysis

Statistical analysis was performed to compare the proportion of patients with lesions in the left and right lungs, as well as in the upper and lower lung lobes of the lungs using the symmetry and homogeneity test for MxN tables with paired samples. A proportion of patients with the presence of lesions on T2 WI and on DWI was compared according to the McNemar criteria. The significance level for all criteria is set to p <0.05.

## 3. Results

The most distinct changes were visualized on T2WI FSE and DWI. On T2WI changes were identified in 9 (60,0%) patients, on DWI – in 5 (33,3%) patients. In 5 (33,3%) patients lesions of the parenchyma were visualized on T2WI and DWI simultaneously. At the same time, 4 (26.7%) patients had changes in lung tissue only on T2WI. (P(McNemar)= 0,125; OR= 0,00 (95%); kappa=0,500) (Table1).

**Table 1 -.** **Comparison of the proportion of patients with the presence of lesions on T2 WI and DWI**

| DWI / T2 WI | No | Yes | Summary |
| --- | --- | --- | --- |
| No | 6 (40,0%) | 4 (26,7%) | 10 (66,7%) |
| Yes | 0 (0,0%) | 5 (33,3%) | 5 (33,3%) |
| Summary | 6 (40,0%) | 9 (60,0%) | 15 (100,0%) |
| P(McNemar) | 0,125 |  |  |
| OR (95% CI) | 0,00 (0,00; 1,51) |  |  |
| kappa | 0,500 |  |  |

Pulmonary lesions had the following distribution in the lungs: in 8 (53,3%) cases, changes were detected in the right lung, in 7 (46,7%) – in the left lung. Comparison of the proportion of patients with a different number of lesions in the left and right lungs: p=0,625 and p=0,31 (Table 2).

**Table 2 -.** **Comparison of the proportion of patients with a different number of lesions in the left and right lung**

| Right /left lung | No | 1-2 lesions | 3 and more lesions | Summary |
| --- | --- | --- | --- | --- |
| No | 6 (40,0%) | 1 (6,7%) | 0 (0,0%) | 7 (46,7%) |
| 1-2 lesions | 2 (13,3%) | 1 (6,7%) | 0 (0,0%) | 3 (20,0%) |
| 3 and more lesions | 0 (0,0%) | 2 (13,3%) | 3 (20,0%) | 5 (33,3%) |
| Summary | 8 (53,3%) | 4 (26,7%) | 3 (20,0%) | 15 (100,0%) |
| P(Symmetry) | 0,625 |  |  |  |
| P (Homogeneity) | 0,311 |  |  |  |

Comparison of the proportion of patients with lesions detected on T2 WI and DWI images showed no significant difference (p = 0.125). The test for symmetry and uniformity also showed no statistically significant difference in the number of lesions in the left and right lungs (p = 0.625 and p = 0.311, respectively). Similar results were obtained when comparing the distribution of lesions between upper and lower lobes of lungs (p = 0.438 and p = 0.405, respectively).

Examples of MRI cases are presented below.

**Case 1:** 45-year-old woman. The clinical manifestations were: fever, cough, chest pain, fatigue. Negative PCR (PCR−).

In basal segments of the lower lobe of the right lung, a section of the inhomogeneously enhanced intensity of the MR signal on T2WI (Fig. 1, a) is identified on the background of the hypointensive lung parenchyma. These changes are less pronounced on Lava Flex Water (Fig. 1, b) and DWI b = 500 sec/mm^2^ (d), probably due to the lower sensitivity of these pulse sequences to vague pulmonary parenchyma edema. IDEAL Water (Fig. 1, c) represents a significantly amplified signal from this section, however, It is extremely difficult to differentiate it with other signal changes due to the low spatial resolution.

**Figure 1.**
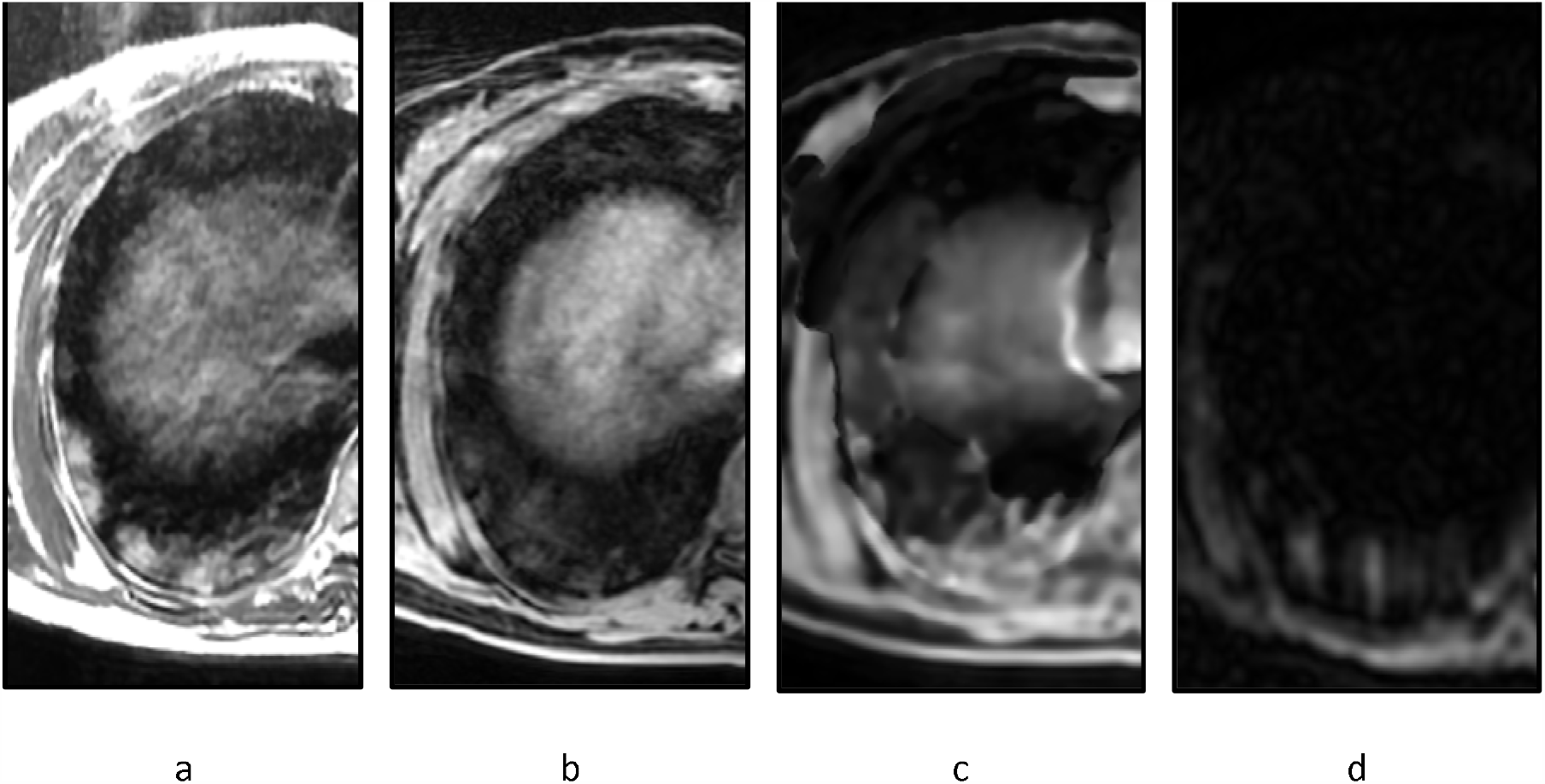
Chest MRI of 45-year-old woman: a – T2 FSE, b – Lava Flex Water, c – IDEAL Water, d – DWI b=500 sec /mm^2^.

**Case 2:** 69-year-old woman. The clinical manifestations were: fever, cough, chest pain, fatigue. Negative PCR (PCR−).

Inhomogeneously enhanced intensity is detected in the basal segments bilaterally on T2WI, which was unclear visualized on Lava Flex Water (Fig. 2, b) on the background of artifacts from chest excursion, while IDEAL Water (Fig. 2, c) shows considerable and extensive changes, which can lead to overdiagnosis of lesions. On DWI b = 500 sec/mm^2^ (Fig. 2, d), right-sided changes are noted, which might be a predictor of consolidation’s development, however, this hypothesis requires confirmation.

**Figure 2.**
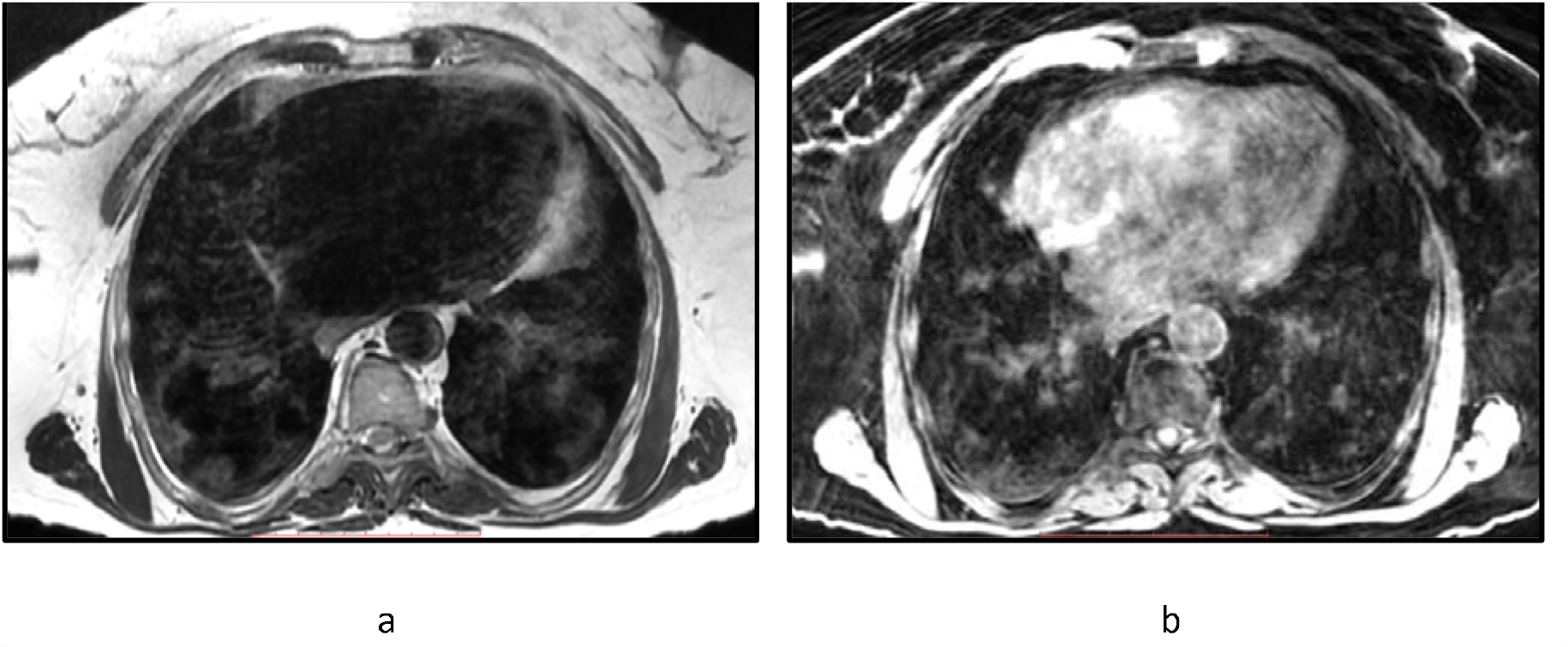
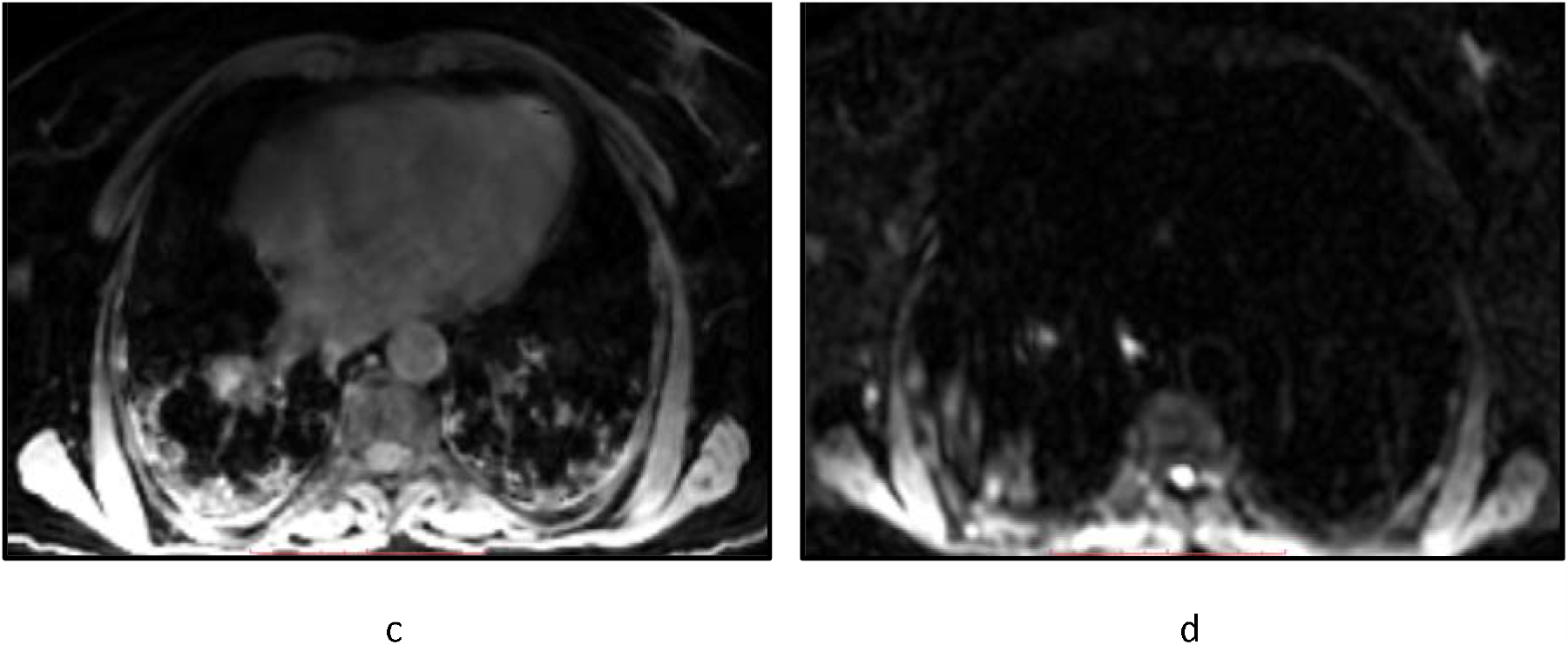
Chest MRI of 69-year-old woman: a – T2 FSE, b – Lava Flex Water, c – IDEAL Water, d – DWI b=500 sec/mm^2^.

**Case 3:** 52-year-old woman. The clinical manifestations were: fever, cough, chest pain, fatigue. Positive PCR (PCR+).

On these MRI tomograms, a pronounced areas of the hyperintensive signal is visualized, both on T2WI (Fig. 3, a), and on all other pulse sequences of Lava Flex Water and IDEAL Water (Fig. b, c). This can indicate a lag in visualizing changes in the lungs to Lava Flex Water.

**Figure 3.**
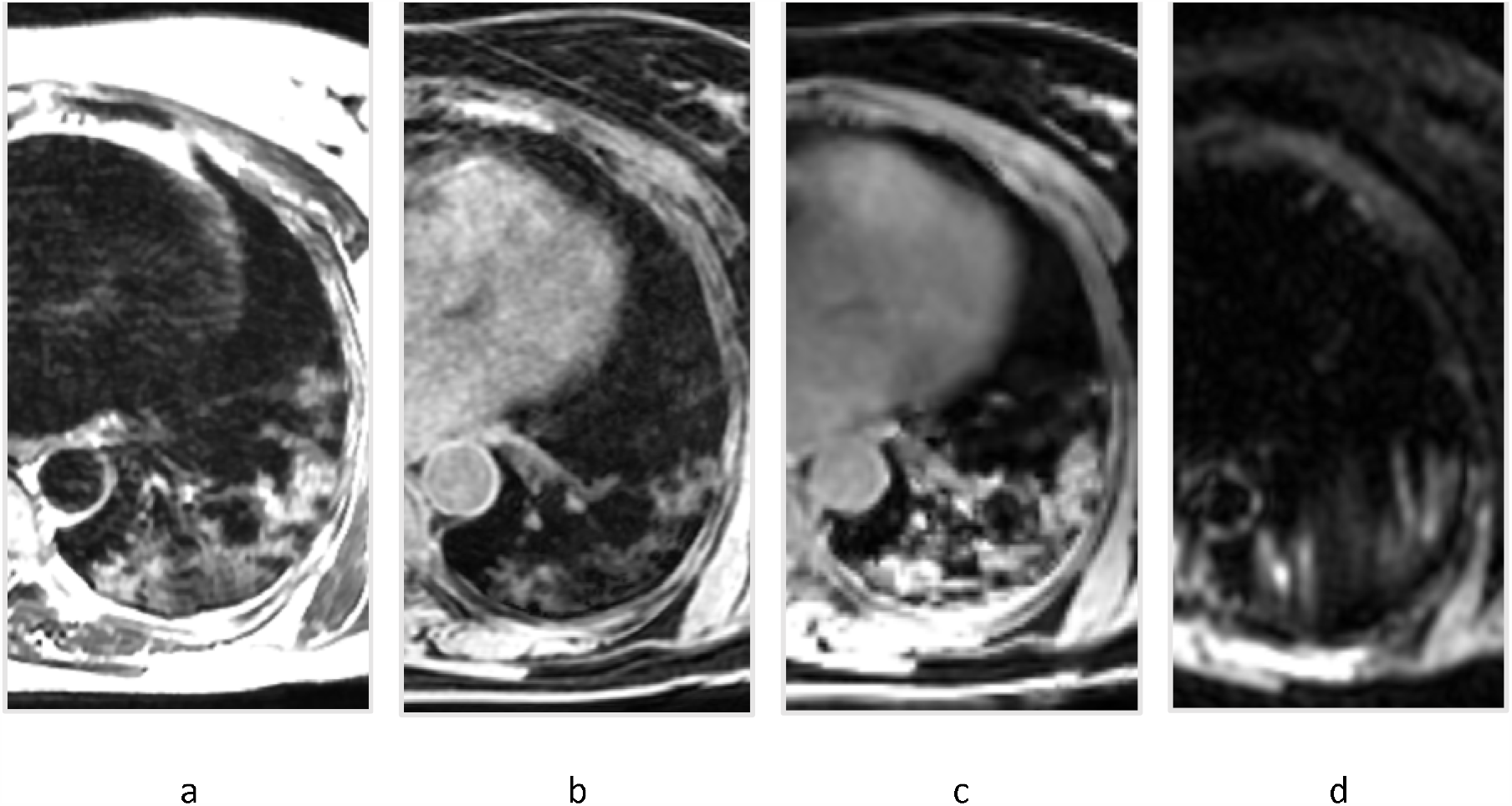
Chest MRI of 52-year-old woman: a – T2 FSE, b – Lava Flex Water, c – IDEAL Water, d – DWI b=500 sec/mm^2^.

**Case 4:** 26-year-old woman. The clinical manifestations were: fever, cough, chest pain, fatigue. Negative PCR (PCR−).

GGOs were detected in S7, 9, 10 (Fig. 4, a) and similar findings were noted in respective segments on all MR-images (Fig. 4, b–f).

**Figure 4.**
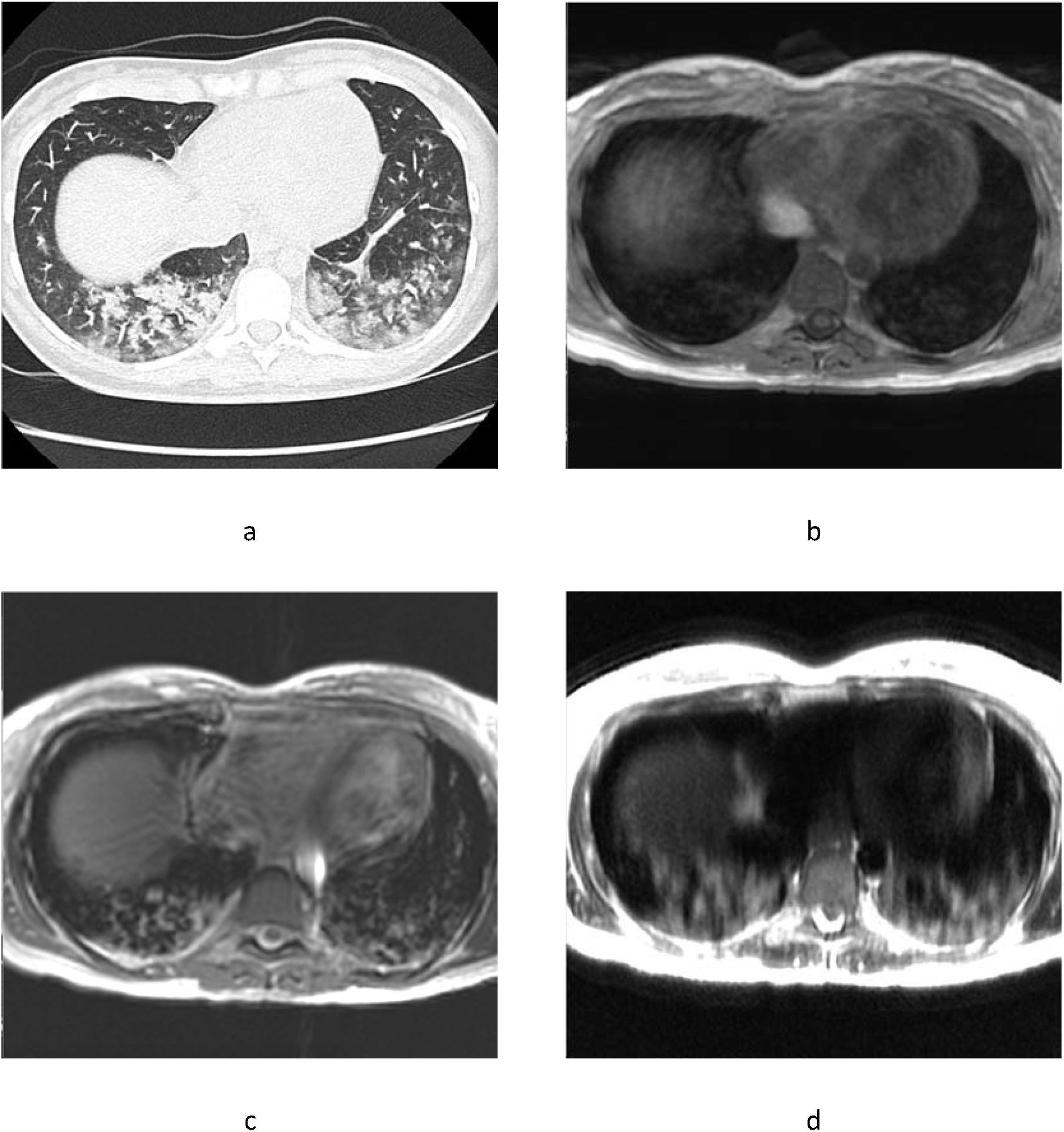
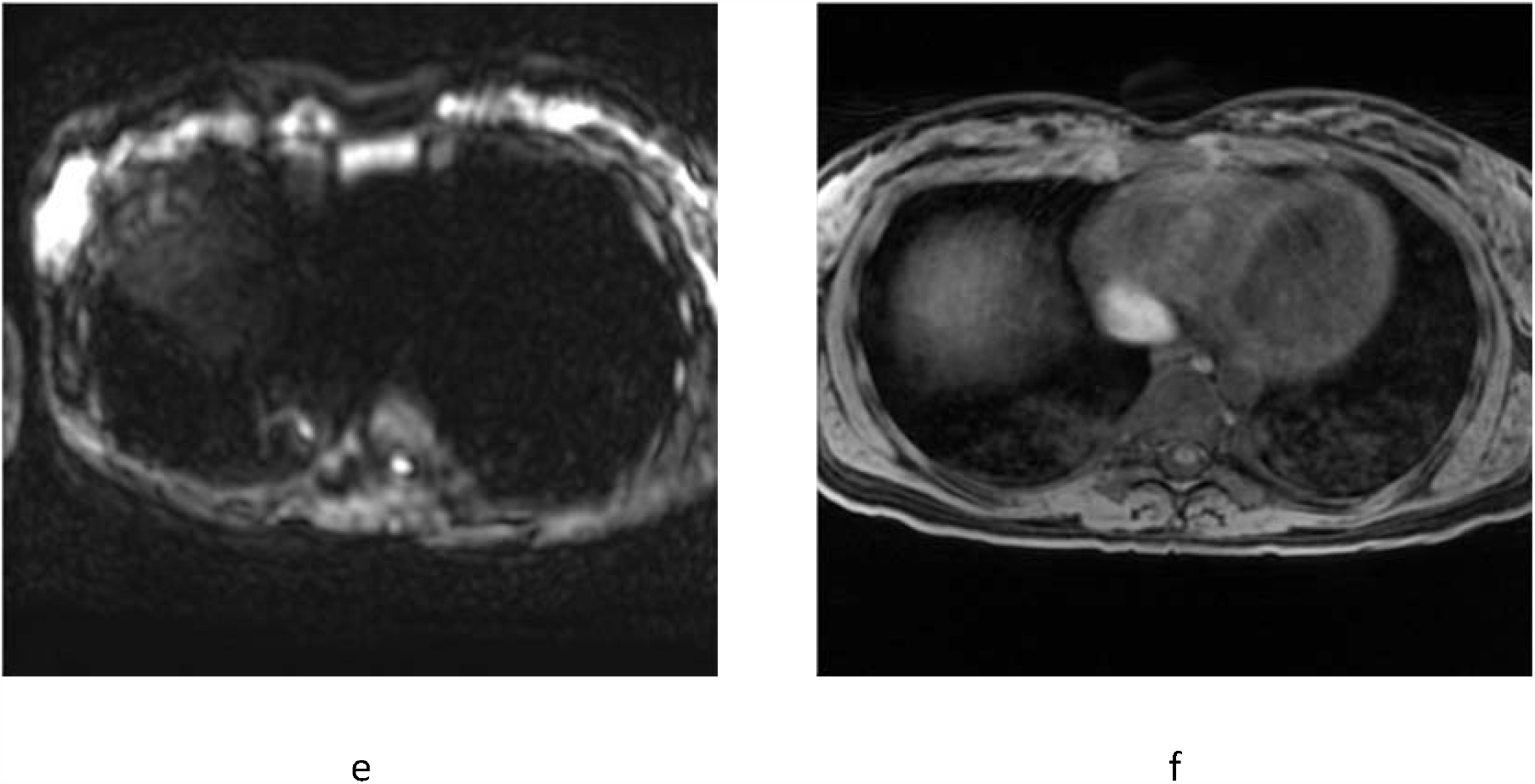
Chest CT scan and MRI of 26-year-old woman: a – axial CT, b – T1WI in-phase, c – T2 fiesta-WI SE, d – T2WI SSFSE, e – DWI (b = 1000 sec/mm^2^), f – T1WI 3D GRE FatSat.

**Case 5:** 26-year-old woman. The clinical manifestations were: fever, cough, chest pain, fatigue. Negative PCR (PCR−).

GGOs were found in S1, 3 of the right lung (Fig. 5, a) and similar findings were noted in respective segments on all MR-images (Fig. 5, b–f).

**Figure 5.**
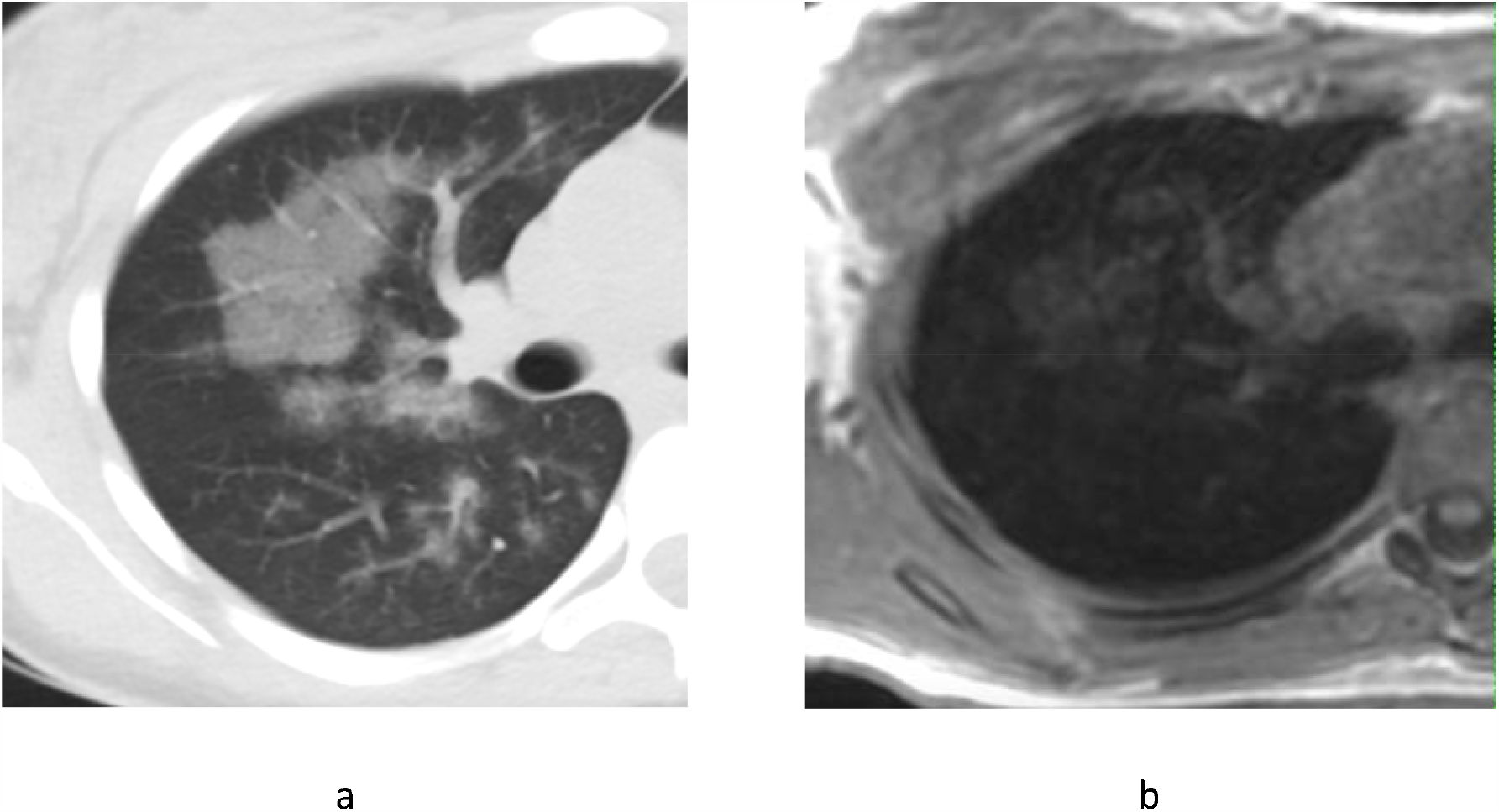
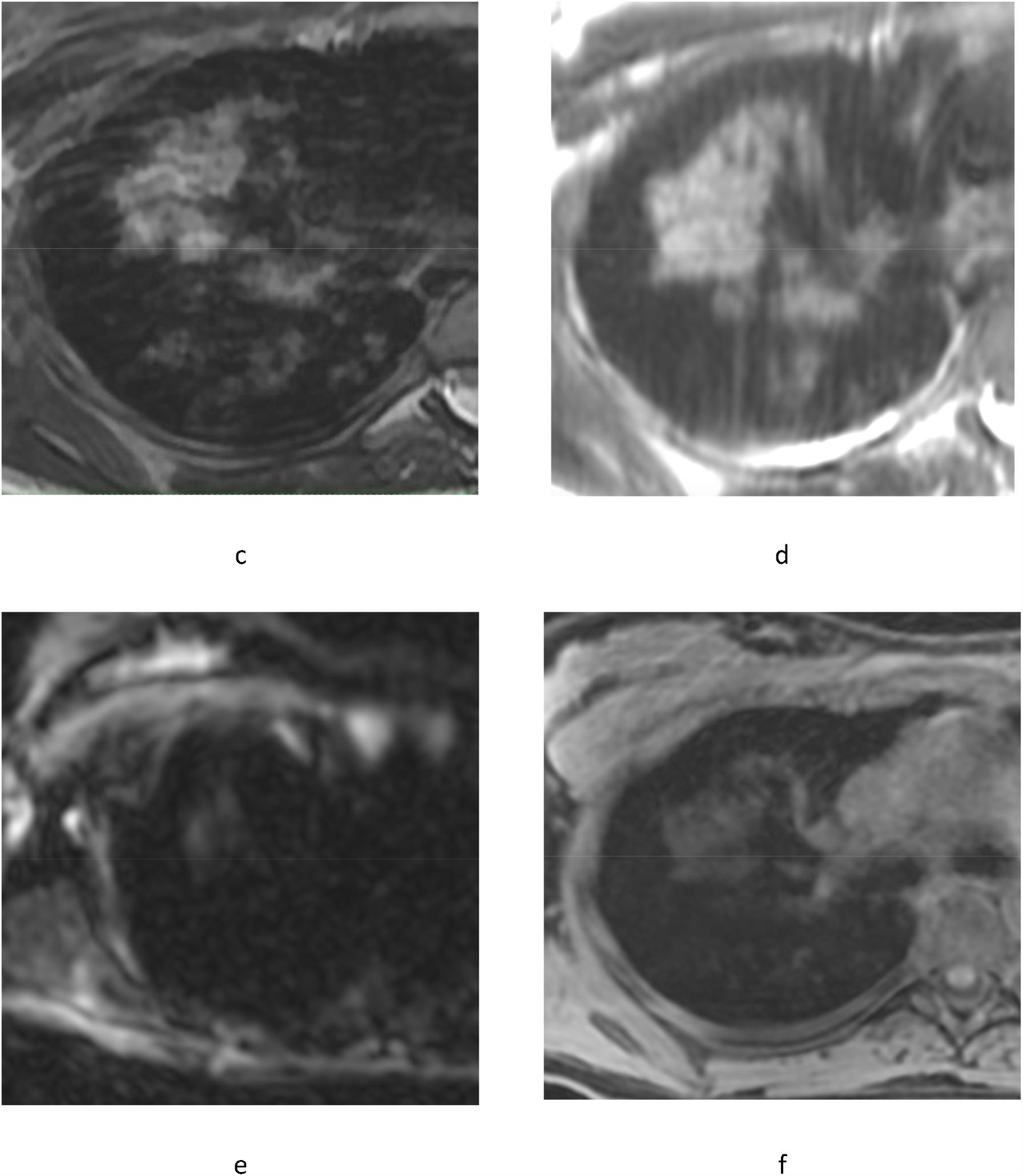
Chest CT and MRI of 26-year-old woman: a – axial CT, b – T1WI in-phase, c – T2 fiesta-WI SE, d – T2WI SSFSE, e – DWI (b = 1000 sec/mm^2^), f – T1WI 3D GRE FatSat.

**Case 6:** 57-year-old man. The clinical manifestations were: fever, cough, chest pain, fatigue. Negative PCR (PCR−).

Consolidation zone visualized on CT scan in S6 of the right lung (Fig. 6, a) is also clearly identif on T2WI with free breathing, but more like GGO (Fig. 6, b). At the same time, the aforemention zone is practically not differentiated on a deep breath (Fig. 6, c).

**Figure 6.**
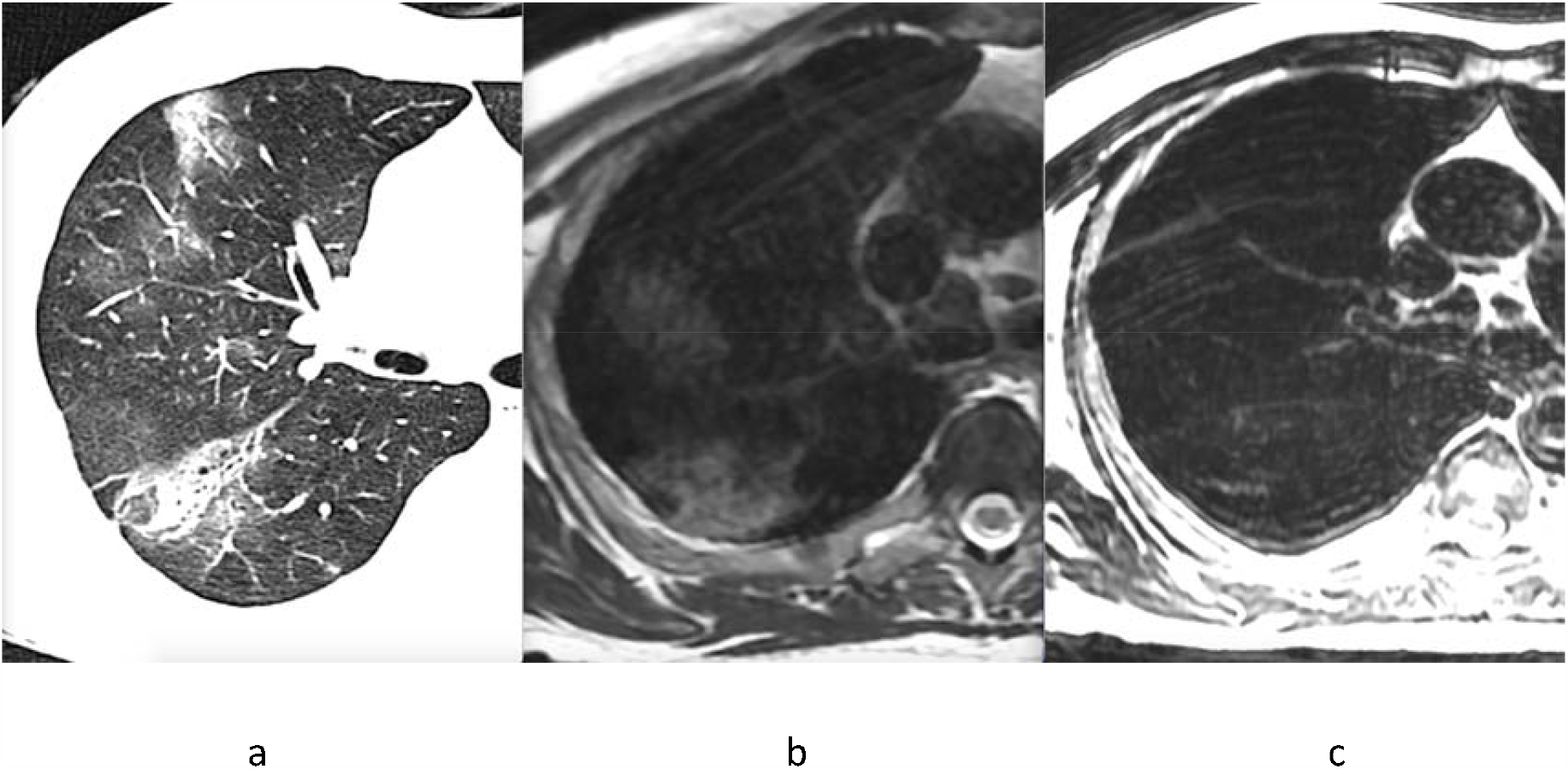
Chest CT and MRI of 57-year-old-man: a – axial CT, b – free breath T2WI, c – deep exhale T2WI.

## 4. Discussion

Although most patients with the novel coronavirus disease (COVID-19) have mild symptoms and good prognosis, in some cases, it is complicated by severe damage to the respiratory system [1]. CT scan is the most sensitive among all imaging modalities for detection of changes in pulmonary parenchyma [2,7]. According to the international expert consensus, CT has become the method of choice for patients with suspected viral pneumonia [3,4,5], since its results directly affect patient management.

Computed tomography plays important role in diagnostics and treatment of lung diseases. Chest CT in patients with COVID-19 pneumonia most commonly demonstrates bilateral, mainly the right lower lobe distribution of subpleural zones of ground glass opacity with fuzzy edges, air bronchograms [6]. Features of viral pneumonia may be presented even in asymptomatic patients, and lesions may quickly increase or consolidate in 1-3 weeks after onset of symptoms, more often at the second week of the disease. Risk factors for worsening prognosis of the disease are old age, male gender, concomitant chronic conditions [7].

Nevertheless, the main question is to what degree detected findings on MRI scans of COVID-19 patients correlate with the radiological changes detected by CT scan. G. Lutterbey et al. showed that high-field MRI has sensitivity comparable to CT scan when diffuse lung changes are detected [11]. At the moment there are no official clinical recommendations or studies comparing the picture of pathological changes in the lungs obtained with CT and MRI at least in small samples of patients with SARS-CoV-2 pneumonia.

The article presents a comprehensive approach to provide highly informative chest MRI as the alternative, non-ionizing method for patients with COVID-19 under the condition of limited availability of computed tomography (CT). In particular, the higher number of averages allowed to increase probability of obtaining high-quality FSE images. At the same time, the refusal to use non-Cartesian filling of k-space, which is often used in practice, allowed us to avoid blurring along images’ periphery. Localization of sections of iso-intense signal was typical for basal and peripheral pulmonary areas, which corresponds to observed changes at CT scan. Crazy-paving signs on chest MRI and CT scans were visually identical. The main weakness of our research is a small sample, which does not allow us to make confident conclusions, as well as to compare the changes we saw on MRI with a possible CT picture of the same patients. The absolute absence of any criteria that would allow us to link the observed MRI picture to severity of the patient’s condition and clinical outcomes complicate the situation.

Magnetic resonance imaging as a method of visualization of the lungs has several disadvantages. Due to the high airiness of the pulmonary tissue, fewer water molecules form a signal, which inevitably reduces the signal-to-noise ratio, resulting in poor-quality images. In addition, susceptibility effects lead to even more pronounced signal attenuation. MRI has low anatomic resolution and is compromised by inevitable artifacts due to breathing movement. However, the ability of MRI to visualize changes in lung structure are constantly expanding due to technical improvement of MRI scanners as well as to appearance of more complex pulse sequences and image post processing [6,7]. Also, MRI with high magnetic field demonstrated a higher signal to noise ratio during lung imaging [8].

In literature, we found works describing clinical cases, where the authors compared CT and MRI [11–16]. Eibel et al. in their study also compared capabilities of high-resolution MRI and CT to detect pulmonary abnormalities suggesting pneumonia in patients with neutropenia. CT and MRI revealed GGOs in 14 and 16 patients, respectively; in one patient, CT scan performed 3 days later showed GGO at the location previously visualized on MRI, indicating sufficient MRI sensitivity to detect GGO [14]. A. Ekinci et al. came to the similar conclusions: all MRI sequences were almost perfectly consistent with detected on CT scans areas of consolidations and local increase in lung density. They believe that MRI can be used, particularly in cases where dynamic monitoring of patients is required in order to avoid ionizing radiation exposure [15]. Consolidation due to infectious pneumonia represented by alveoli filled with fluid, gives a high signal intensity on T2WI. MRI can also differentiate the consolidation associated with fibrous tissues by the relatively “short” T2 component. It can be easily distinguished from a focal consolidation associated with pulmonary infarction, which is caused by interalveolar blood. In this case, increase in signal intensity on T1WI will be observed due to the formation of methemoglobin during subacute hemorrhage [16].

## 5. Conclusion

The results showed that in case of CT scan is not available, it is advisable to conduct a chest MRI for patients with suspected or confirmed COVID-19. It is important that in contrast to planar imaging methods, MRI allows us to assess the extent of the lesion and monitor the disease dynamics. It should be noted that the formations on MRI do not look like GGO, therefore in this work we call them “cloudy sky sign”.

These data also may be useful in interpreting other studies, such as thoracic spine MRI, cardiac MRI, detecting signs of viral pneumonia in asymptomatic individuals.

Currently, we have registered an observational prospective study (ClinicalTrials.gov Identifier: NCT04424355), which plans to determine MRI capabilities in diagnostics of viral pneumonia in a large sample of patients.

## Data Availability

Data is no available

## Ethics approval

The study was approved by Local Ethics Committee of Morozov Children Municipal University Hospital of the Moscow Healthcare Department.

